# Diagnostic performance of plasma Aβ_1-42_, Aβ_1-40_ and pTau_181_ in the LUMIPULSE automated platform for the detection of Alzheimer disease

**DOI:** 10.1101/2023.04.20.23288852

**Authors:** Javier Arranz, Nuole Zhu, Sara Rubio-Guerra, íñigo Rodríguez-Baz, María Carmona-Iragui, Isabel Barroeta, Ignacio Illán-Gala, Miguel Santos-Santos, Juan Fortea, Alberto Lleó, Mireia Tondo, Daniel Alcolea

## Abstract

**BACKGROUND:** Recently-developed blood markers for Alzheimer’s (AD) detection have high accuracy but usually require ultra-sensitive analytic tools not commonly available in clinical laboratories.

**METHODS:** We analyzed plasma samples from 367 consecutive participants in the SPIN cohort, comprising 302 euploid participants (67 cognitively unimpaired, 136 participants with mild cognitive impairment, and 99 with dementia) and 65 with Down Syndrome (46 non-demented and 19 with AD dementia). Participants were classified according to CSF biomarkers status using the AT(N) system. Plasma Aβ_1–42_, Aβ_1–40_ and pTau_181_ were measured in the fully-automated LUMIPULSE platform. We used ANOVA to compare plasma biomarkers concentrations between AT(N) groups, evaluated Spearman’s correlation between plasma and CSF and performed ROC analyses to assess their diagnostic accuracy to detect AD.

**RESULTS:** Plasma pTau_181_ concentration was higher in A+T+ than A+T- and A-T-, and in A+T- and A-T+ than A-T-. The plasma Aβ_1–42_/Aβ_1–40_ ratio was lower in A+T+ and A+T- compared to A-T-. pTau_181_ and the Aβ_1–42_/Aβ_1–40_ ratio showed moderate correlation between plasma and CSF (Rho=0.66 and 0.65, respectively). The areas under the ROC curve (AUC) to discriminate A+T+ from A-T- participants were 0.91 for pTau_181_ and 0.86 for Aβ_1–42_/Aβ_1–40_. The combination of both measures yielded an AUC=0.94. Chronic kidney disease (CKD) was related to increased plasma biomarker concentrations, but ratios were not significantly affected.

**CONCLUSION:** The feasibility and performance of plasma-based biomarker measurements on an automated platform showed high diagnostic accuracy and hold great promise for the diagnostic process of AD.

**What is already known on this topic:** Blood biomarkers have shown high accuracy to detect AD pathophysiology. The feasibility of those biomarkers in different platforms and the influence of comorbidities in their concentrations needs to be studied.

**What this study adds:** We analyze the feasibility and diagnostic performance of AD biomarkers measured in a fully-automated platform and assess how comorbidities affect their concentrations.

**How this study might affect research, practice, or policy:** The measurement of plasma AD biomarkers in an automated platform yields high accuracy to detect AD pathophysiology and would be easy to implement. Plasma AD biomarker concentrations are increased in chronic kidney disease, and in this context, the use of ratios would be more reliable.

## Introduction

Early and accurate diagnosis is becoming an increasing priority with the recent developments of disease-modifying therapies for Alzheimer’s Disease (AD). Pathophysiological biomarkers in cerebrospinal fluid (CSF) and positron emission tomography (PET) imaging with amyloid and tau tracers have extensively proven to be useful to detect the presence of the disease but are either expensive and/or invasive [1], which can delay the diagnosis and access to a treatment.

The measure of AD biomarkers in blood through reliable high-throughput platforms would simplify the diagnostic process. This is now technically possible thanks to the development of sensitive technologies that can consistently quantify brain-derived molecules that are present in blood in very low concentrations[2–4]. Amyloid-β (Aβ) peptides and different isoforms of phosphorylated tau (pTau) in blood have shown high accuracy to detect AD pathophysiology in previous research studies [5–11]. How these plasma markers are affected by different comorbidities is also starting to be understood thanks to large well-characterized cohorts [12–14]. Thus, blood-based markers have the potential to be of great use in the screening, early diagnosis, tracking progression, and ultimately, monitoring the efficacy of treatment [15–17]. However, most of the existing studies have assessed the value of these markers individually or through techniques not widely available in laboratories, limiting their potential to be widely applied in the clinical routine. Their implementation of blood AD markers in a fully-automated platform would facilitate their reproducibility and accessibility in clinical laboratories [18].

The fully-automated platform LUMIPULSE G, extensively used to measure CSF AD biomarkers, has recently launched specific assays to measure Aβ_1-42_, Aβ_1-40_ and pTau_181_ in plasma. In this study, our aim was to assess the feasibility and diagnostic performance of Aβ_1-42/_Aβ_1-40_ ratio and pTau_181_ in plasma in the LUMIPULSE fully-automated platform in a retrospectively well characterized cohort of individuals.

## Methods

### Study participants and clinical classification

We included consecutive individuals who underwent lumbar puncture for the analysis of AD CSF biomarkers assessed at the Sant Pau Memory Unit (Barcelona, Spain) as part of the SPIN cohort [19] between January 2021 and December 2021. The study was approved by the Sant Pau Ethics Committee (Protocol code: EC/22/202/6880) following the standards for medical research in humans recommended by the Declaration of Helsinki. All participants or their legally authorized representative gave written informed consent to participate in biomarkers research studies.

Participants had a diagnosis of dementia, mild cognitive impairment (MCI), or were cognitively unimpaired (CU). The clinical diagnosis was established after a thorough neurological and neuropsychological evaluation[19]. A subset of participants had Down syndrome and were evaluated in the context of the Down Alzheimer Barcelona Neuroimaging Initiative (DABNI) linked to a population-based health plan in Catalonia, Spain, run at the Barcelona Down Medical Center[20]. For Down syndrome participants, diagnosis of dementia was based on a neurological and neuropsychological examination that included semistructured health questionnaires and a neuropsychological battery adapted for intellectual disabilities [21]. The subset of participants with Down syndrome were classified clinically into 2 groups in a consensus meeting between the neurologist and neuropsychologist after independent visits: without dementia, that included asymptomatic and prodromal AD, and AD with dementia. Participants were classified according to the estimated glomerular filtrate rate (eGFR) in different stages of chronic kidney disease (CKD).

After a full evaluation that included analysis of AD CSF biomarkers, participants were classified according their etiologic diagnosis, as AD with pathophysiological evidence (AD), other neurodegenerative dementias (OtherDem) or CU. A proportion of participants’ diagnosis was classified as “uncertain” as they had an unclear etiological diagnosis after a full initial evaluation and required clinical follow-up.

### Sample collection and analysis

Blood samples were collected in EDTA-K2 tubes and subsequently centrifuged (2000rpm x 10 mins, 4ºC) within 2 hours after extraction. Plasma was aliquoted and stored at -80ºC until analysis. CSF samples were obtained through lumbar puncture, and were also centrifuged, aliquoted and stored at -80ºC until analysis. Full protocol for CSF sample collection in our center has been previously reported[19].

All samples were measured in the Lumipulse fully-automated platform G600II using commercially available kits (Fujirebio Europe, Ghent, Belgium) for Aβ_1–42_, Aβ_1–40_, and pTau_181_. Plasma samples were analyzed between July and August 2022 with the same lot of reagents. On the day of the analysis, plasma samples were brought to room temperature, mixed thoroughly, and centrifuged for 5 minutes at 2000g, and subsequently transferred to specific cuvettes for analysis in the Lumipulse platform.

CSF markers Aβ_1–42_, Aβ_1–40_, pTau_181_ and tTau were used in the diagnostic assessment of patients and measured in routine runs scheduled twice a month throughout 2021 following previously reported methods[22]. According to CSF markers, all participants were classified as amyloid positive (A+, CSF Aβ_1–42_/Aβ_1–40_<0.062) or negative (A-), and as tau positive (T+, pTau_181_ >63pg/mL) or negative (T-). Validation of these cutoff values has been described elsewhere[22].

DNA was extracted from full blood using standard procedures, and *APOE* was genotyped following previously reported methods[19]. Briefly, direct DNA sequencing of exon 4 was performed routinely for all participants in the SPIN cohort, followed by visual analysis of the resulting electropherogram to identify the two coding polymorphisms that encode the three possible apoE isoforms.

### Statistical analysis

Data normality was assessed with the Shapiro-Wilk test. Non-normally distributed variables were log-transformed when necessary. ANCOVA test adjusted by age and sex followed by Tukey’s post hoc correction test was performed. To assess differences in categorical variables, Chi Square test was used. To assess the correlation between plasma and CSF markers, Spearman test was used.

Diagnostic accuracy of plasma biomarkers was assessed through receiver operating characteristic (ROC) analysis. The areas under the curve (AUC) of individual markers were calculated and logistic regression models that combined them with each other together with clinical variables were performed. Model 0 included Age, Sex and *APOE4* status. Model 1 included pTau_181_ and Aβ_1–42/_ Aβ_1-40_. Model 2 included pTau_181_ and Aβ_1– 42_, and model 3 included Age, Sex, *APOE4*, and pTau_181_, Aβ_1–42_ and Aβ_1–40_.We compared their accuracy using DeLong’s test. We calculated a range of plasma cutoffs and their sensitivity, specificity and Youden’s J index to discriminate A+T+ from A-T-. We also provide plasma cut-offs that maximized the Youden’s J index and those that yielded 95% sensitivity (optimized for screening purposes). All tests were performed in R statistical software version 4.2.1. Alpha threshold was set at 0.05 for all analysis.

## Results

### Study participants and clinical classification

We included 302 euploid participants previously classified as cognitively unimpaired (CU, n=67), with mild cognitive impairment (MCI, n=136) or with dementia (n=99). We also included 65 participants with Down syndrome (46 without dementia and 19 with dementia). **Table 1** shows the main demographic characteristics and biomarker measures in each group. CU participants were younger than those with MCI (p<0.001) and those with dementia (p<0.001) groups. As expected, Down Syndrome participants without dementia were younger than those with dementia (p=0.008). There were more female participants in the euploid group (61.6%) and more male participants in the Down syndrome group (61.5%). The proportion of A+T+ and *APOE4* positive increased according to the clinical stage.

**Table 1:** Demographics and biomarker concentrations in CSF and plasma. Unless otherwise specified, values are presented as mean (SD). CU: Cognitively Unimpaired. AD: Alzheimer disease. OtherNotDeg: Other not neurodegenerative. OtherDem: Other dementias. MCI: Mild cognitive impairment. DS.NotDem: Down Syndrome without dementia. DS.Dementia: Down Syndrome with dementia.

|  | Euploid |  |  | Down syndrome |  |  |
| --- | --- | --- | --- | --- | --- | --- |
|  | CU | MCI | Dementia | DS.NotDem | DS.Dementia | P-value |
| n | 67 | 136 | 99 | 46 | 19 |  |
| Age (years) | 58.1 (12.1) | 71.4 (8.61) | 73.4 (7.97) | 43 (11.9) | 51.5 (3.46) | <0.001 |
| MMSE score | 29.1 (0.947) | 25.4 (3.17) | 21.5 (4.19) | NA | NA | <0.001 |
| Female (%) | 70.1 | 57.4 | 61.6 | 41.3 | 31.6 | 0.0013 |
| *APOE4*+ (%) | 19.4 | 23.7 | 38.4 | 21.7 | 31.6 | <0.001 |
| A+T+ (%) | 4.48 | 36.8 | 57.6 | 37 | 78.9 | <0.001 |
| CSF Aβ<sub>1-42</sub> (pg/mL) | 1039 (400) | 791 (410) | 593 (262) | 757 (384) | 498 (179) | <0.001 |
| CSF Aβ<sub>1-40</sub> (pg/mL) | 10501<br>(3436) | 11323<br>(4276) | 10380<br>(3630) | 11754<br>(5240) | 11975 (6097) | ns |
| CSF Aβ<sub>1-42</sub>/Aβ<sub>1-40</sub> | 0.099<br>(0.017) | 0.072<br>(0.0271) | 0.06<br>(0.0223) | 0.067<br>(0.025) | 0.044<br>(0.00858) | <0.001 |
| CSF tTau (pg/mL) | 292 (129) | 490 (316) | 614 (389) | 455 (343) | 975 (711) | <0.001 |
| CSF pTau<sub>181</sub> (pg/mL) | 36 (17.6) | 73.7 (55.9) | 94.3 (71.7) | 70.9 (67.5) | 176 (169) | <0.001 |
| plasma pTau<sub>181</sub><br>(pg/mL) | 1.84<br>(0.606) | 2.52<br>(0.968) | 3.43 (1.52) | 2.92 (1.56) | 5.75 (2.23) | <0.001 |
| plasma Aβ<sub>1-42</sub><br>(pg/mL) | 25 (3.65) | 24.4 (6.35) | 25.1 (5.9) | 36.9 (5.27) | 36.8 (5.72) | <0.001 |
| plasma Aβ<sub>1-40</sub><br>(pg/mL) | 295 (44.3) | 316 (68.6) | 335 (70.9) | 464 (65.7) | 496 (73.1) | <0.001 |
| plasma Aβ<sub>1-42</sub>/Aβ<sub>1-40</sub> | 0.085<br>(0.008) | 0.077<br>(0.01) | 0.075<br>(0.008) | 0.08<br>(0.011) | 0.074 (0.006) | <0.001 |
| plasma pTau<sub>181</sub>/Aβ<sub>1-42</sub> | 0.074<br>(0.0244) | 0.108<br>(0.046) | 0.143<br>(0.085) | 0.081<br>(0.044) | 0.157 (0.058) | <0.001 |
| CU/AD/OtherNotDeg/<br>OtherDem/Down/<br>Uncertain | 57/4/4/0/0/<br>2 | 0/43/46/9/0<br>/38 | 0/50/11/23/0/<br>15 | 0/0/0/0/46/0 | 0/0/0/0/19/0 |  |

### Measures of Aβ_1–42_, Aβ_1–40_ and pTau_181_ in plasma

All plasma measures for Aβ_1–42_, Aβ_1–40_ and pTau_181_ were above their lower limit of quantification. The plasma concentrations in the study ranged from 14.06 to 65.65 pg/mL for Aβ_1–42_, 180.41 to 780.44 pg/mL for Aβ_1–40_ and 0.92 to 10.32 pg/mL for pTau_181_. Inter-assay coefficients of variation were assessed at two levels (low and high concentrations) for each analyte and were 5.7% (21 pg/mL) and 6.6% (209 pg/mL) for Aβ_1–42_, 5.5% (214 pg/mL) and 6.7% (2243 pg/mL) for Aβ_1–40_, and 4.6% (5 pg/mL) and 4.1% (45 pg/mL) for pTau_181_.

### Correlation between plasma and CSF biomarkers

As per inclusion criteria, all participants had CSF biomarkers measures. We explored the correlation between both matrices. The correlation between plasma and CSF was moderate for pTau_181_ (Rho = 0.66, p<0.001) and low for Aβ_1–42_ (Rho = 0.14, p= 0.007), and Aβ_1–40_ (Rho 0.1, p= 0.048). When using ratios, the correlation was high for pTau_181_/Aβ_1–42_ (Rho 0.79, p< 0.001), and moderate for Aβ_1–42_/Aβ_1–40_ (Rho= 0.65, p<0.001). Detailed correlations within clinical subgroups are shown in **Supplementary Material**.

### Association between plasma biomarkers and AT status in CSF

We assessed the differences between distinct AT status considering other variables in a multivariate model. We studied the effect of age, sex, *APOE* status (*APOE4*+), renal function measured by the estimated glomerular filtration rate (eGFR), vascular risk factors (presence of at least one of the following: high blood pressure, diabetes mellitus, dyslipidemia, history of stroke, obstructive sleep apnea with CPAP) and clinical status (CU, MCI and Dementia, and Down Syndrome with and without dementia). As shown in **Figure 1**, the multivariate model confirmed that the A+T+ group had higher plasma concentrations of pTau_181_ compared to A+T- (p=0.0014), A- T+ (p=0.0008) and A-T- (p<0.0001) groups. The A+T- group also had higher concentrations of pTau181 compared to A-T- group (p<0.0001). In turn, the A+T+ group had lower levels of Aβ_1–42_ compared to the A-T+ (p=0.034) and the A-T- (p<0.001) groups. Similar results were seen using the ratios pTau_181_/Aβ_1–42_ and Aβ_1– 42_/Aβ_1–40._ The plasma pTau_181_/Aβ_1–42_ ratio was higher in A+T+ compared to A-T- (p<0.001), A-T+ (p<0.001) and A+T- (p=0.001) groups. It was also higher in A+T- compared to A-T- (p<0.001) and A-T+ (p= 0.04) groups. The plasma Aβ_1–42_/Aβ_1–40_ ratio was lower in A+T+ and A+T- compared to A-T- (both p<0.001).

**Figure 1.**
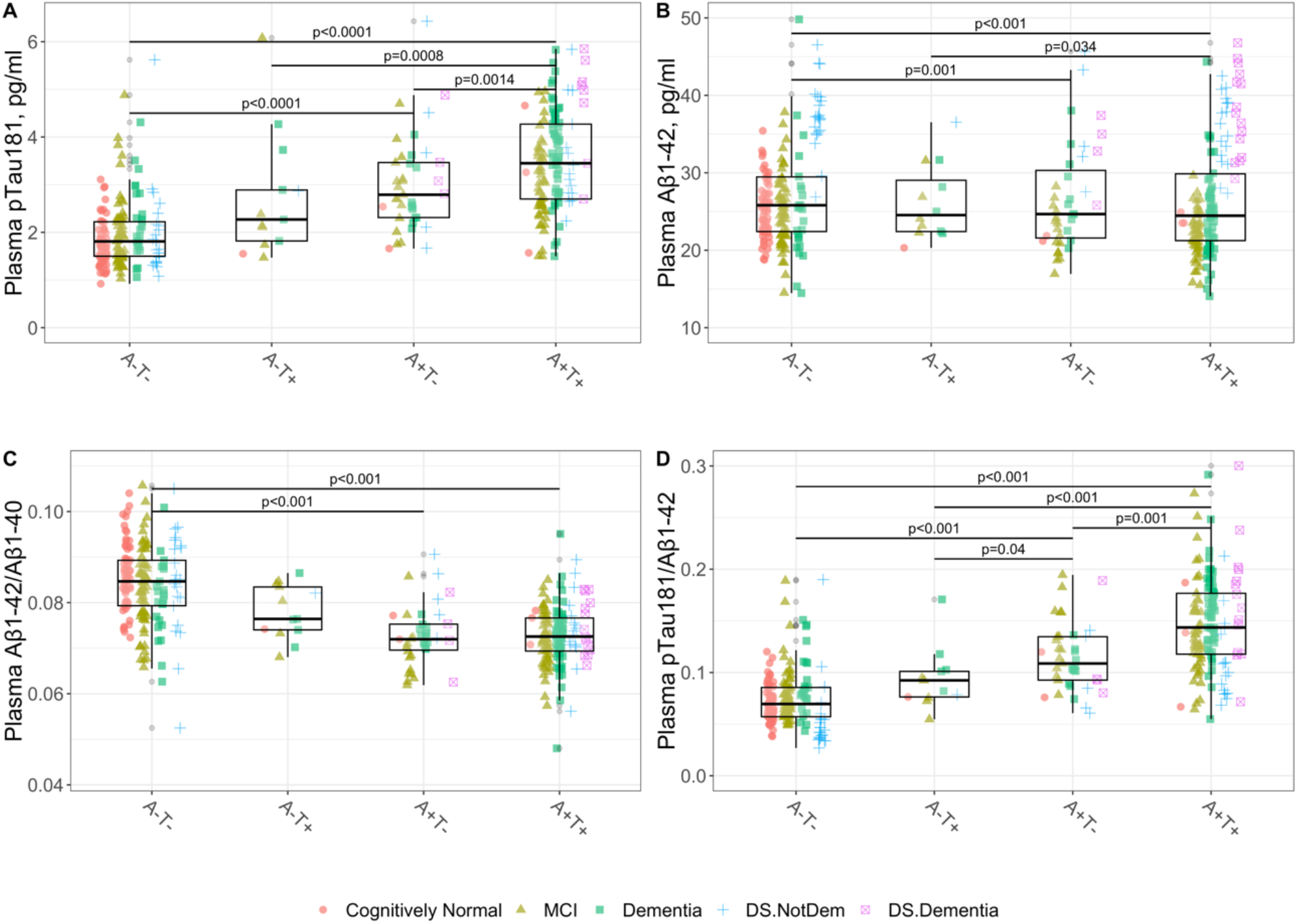
Levels of plasma biomarkers and their ratios according to the AT status in CSF. All p-values are derived from multivariate linear model, adjusted for the effects of age, sex, *APOE4* status, chronic kidney disease stage, vascular risk factors and clinical stage. pTau_181_: phosphorylated tau 181. Aβ_1–42_: Amyloid β_1–42_. Aβ_1–40_: Amyloid β_1–40_. MCI: Mild cognitive impairment. DS.NotDem: Down Syndrome without dementia. DS.Dementia: Down Syndrome with dementia.

### Effect of other variables on plasma biomarkers

We assessed whether plasma pTau_181_ and Aβ_1–42_/Aβ_1–40_ were affected by other variables in the multivariate model. **Figure 2** shows the effect of each variable represented by the standardized beta coefficient. We observed that the amyloid positivity (A+T- and A+T+) and Down syndrome were associated to higher plasma concentration of pTau_181_ and lower Aβ_1–42_/Aβ_1–40_ ratio. Decreased renal function was associated with higher concentrations of pTau_181_ and higher Aβ_1–42_/Aβ_1–40_ ratio. Male sex was associated with higher pTau181. The Aβ_1–42_/Aβ_1–40_ ratio was lower in the dementia group. Our model had and adjusted R^2^ value of 0.6 for pTau_181_ and 0.41 for Aβ_1–42_/Aβ_1–40_.

**Figure 2.**
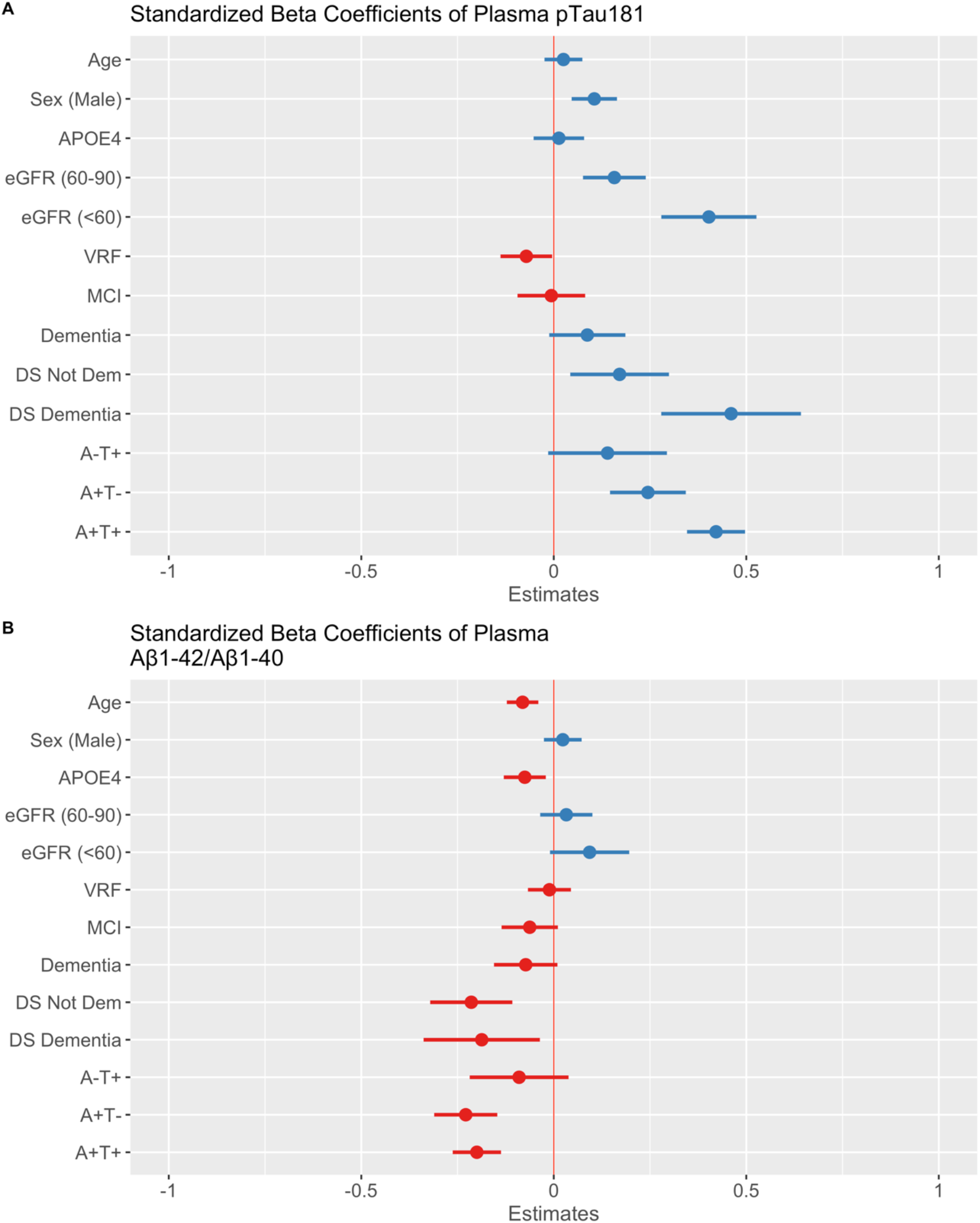
Effect of different variables on plasma pTau_181_ and Aβ_1–42_/Aβ_1–40_. Dots and bars represent the standardized beta coefficients of each variable in a multivariate regression model. Lines represent the 95% confidence interval for each standardized beta coefficient. Red vertical dashed lines indicate a null effect. In red the negative standardized beta coefficients and in blue the positive standardized beta coefficients. pTau_181_: phosphorylated tau 181. MCI: mild cognitive impairment. DS: Down Syndrome. VRF: vascular risk factors. eGFR: estimated glomerular filtration rate.

As impairment of renal function had a significant effect on plasma markers, we performed a subanalysis after stratifying by eGFR. We found that pTau_181_ concentration in plasma was higher as renal function decreased (<60 vs. >60 mL/min/1.73m^2^ and 60-90 vs. >90 mL/min/1.73m^2^, p<0.001), Aβ_1–42_ and Aβ_1–40_ concentrations in plasma were also higher as renal function decreased (p<0.001), but those differences were lost when using the Aβ_1–42_/Aβ_1–40_ or the pTau_181_/ Aβ_1–42_ ratios.

### Diagnostic accuracy of plasma biomarkers and their combinations for the discrimination of A+T+ from A-T-

When we compared accuracies with DeLong test, we adjusted p-value by multiple comparisons. The AUC to discriminate A+T+ from A-T- participants 0.91 (CI 0.87-0.94) for pTau_181_ and 0.86 (CI 0.82-0.91) for Aβ_1–42_/Aβ_1–40_ (**Figure 3**). These accuracies were significantly higher than those from a basic model that included age, sex and *APOE4* status (p<0.001). Aβ_1–42_ and Aβ_1–40_ individually had poor diagnostic accuracy, yielding AUCs below 0.70.

**Figure 3.**
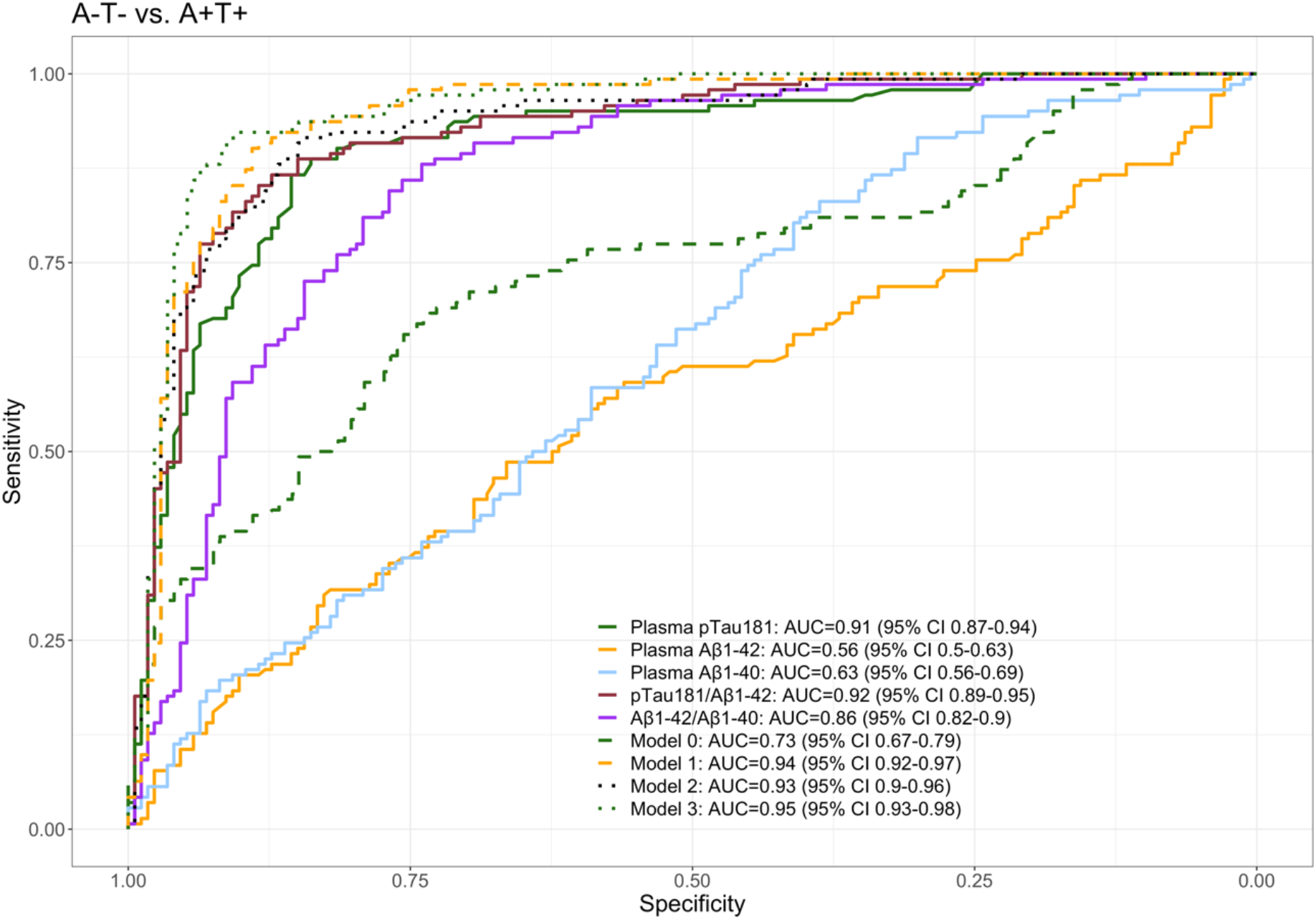
Diagnostic accuracy of plasma biomarkers for the discrimination of A+T+ from A-T- categories. pTau_181_: phosphorylated tau 181. Aβ_1–42_: Amyloid β_1–42_. Aβ_1–40_: Amyloid β_1–40_. Model 0: Age+Sex+*APOE4*. Model 1: pTau_181_ + Aβ_1–42/_ Aβ _1-40_. Model 2: pTau_181_ + Aβ_1–42_, Model 3: Age+Sex+*APOE4*+pTau_181_+ Aβ_1–42_+ Aβ_1–40_.

Diagnostic accuracies of pTau_181_/Aβ_1–42_, Aβ_1–42_/Aβ_1–40_ and a logistic regression model combining pTau_181_ and Aβ_1–42_ (model 2) were not significantly higher than that of pTau_181_ alone (p= 0.18, p=0.076 and p=0.007, respectively, not significant adjusted by multiple comparisons). The combination of pTau_181_ and Aβ_1–42_/Aβ_1–40_ (model 1) yielded an AUC of 0.94 (CI 0.92-0.97), which was higher than that of Aβ_1–42_/Aβ_1–40_ (p < 0.0001) but not compared to pTau_181_ (p=0.0025, not significant adjusted by multiple comparisons). The addition of other variables (age, sex, *APOE4* status) to pTau_181_ did not significantly increase its accuracy. Detailed two-by-two comparisons can be found as **Supplementary Material**. We obtained similar results when the outcome was restricted to discriminate A+T+ patients with a clinical diagnosis of AD from A-T- patients with “Other dementias”, A-T- patients with “Other not neurodegenerative” cognitive impairment, and from A-T- CU participants. We also assessed the performance of the biomarkers in intermediate AT states (A-T+ and A+T-) (**Supplementary Material**).

### Cutoffs application

**Table 2** shows the accuracy of different thresholds for pTau_181_ and for Aβ_1–42_/Aβ_1–40_ to detect A+T+ participants. For pTau_181_, a cutoff value of 2.4 pg/mL yielded the highest Youden J index with a sensitivity of 89% and a specificity of 84%. Considering an intended use for screening, we also calculated the cutoff value that maximized sensitivity to 95%. A cutoff value of 2.01 pg/mL, with a sensitivity of 95%, yielded a specificity of 65%. The use of this cutoff value in our cohort resulted in 78% correctly classified individuals (135 true positive, 112 true negative), and 22% misclassified individuals (7 false negative, 61 false positive). However, the sequential application of the plasma Aβ_1–42_/Aβ_1–40_ ratio (cutoff 0.083) in participants that were pTau_181_ negative reduced the number of false negative to 0 without increasing the number of false positive in the whole cohort and in all the subsets analyzed (**Table 2**). These findings suggest that an algorithm that considers the sequential application of plasma markers could be valuable for a more accurate detection of the AD pathology (**Figure 4)**.

**Table 2.** Thresholds for plasma pTau_181_, Aβ_1–42_/Aβ_1–40_ and pTau_181_/ Aβ_1–42_ ratios to detect A+T+ participants. Shaded cells indicate cutoffs that yielded the maximum Youden J indices and those with 95% sensitivity.

| Threshold pTau<sub>181</sub> | Sensitivity | Specificity | Youden |
| --- | --- | --- | --- |
| 1.51 | 0.98 | 0.25 | 0.233 |
| 2.01 | 0.95 | 0.65 | 0.598 |
| 2.34 | 0.90 | 0.82 | 0.716 |
| 2.43 | 0.89 | 0.84 | 0.725 |
| 2.56 | 0.85 | 0.86 | 0.708 |
| 2.82 | 0.74 | 0.90 | 0.635 |
| 3.34 | 0.58 | 0.95 | 0.525 |
| 4.91 | 0.20 | 0.99 | 0.186 |
| Threshold Aβ<sub>1-42</sub>/ Aβ<sub>1-40</sub> | Sensitivity | Specificity | Youden |
| 0.087 | 0.99 | 0.38 | 0.367 |
| 0.083 | 0.95 | 0.57 | 0.517 |
| 0.081 | 0.90 | 0.69 | 0.595 |

| 0.080 | 0.88 | 0.74 | 0.620 |
| --- | --- | --- | --- |
| 0.078 | 0.85 | 0.77 | 0.614 |
| 0.075 | 0.68 | 0.85 | 0.526 |
| 0.074 | 0.59 | 0.90 | 0.488 |
| 0.068 | 0.18 | 0.96 | 0.143 |
| 0.066 | 0.09 | 0.99 | 0.080 |
| Threshold pTau<sub>181</sub>/Aβ<sub>1-42</sub> | Sensitivity | Specificity | Youden |
| 0.067 | 0.99 | 0.46 | 0.448 |
| 0.075 | 0.95 | 0.61 | 0.558 |
| 0.090 | 0.90 | 0.81 | 0.711 |
| 0.099 | 0.87 | 0.87 | 0.739 |
| 0.102 | 0.85 | 0.88 | 0.729 |
| 0.106 | 0.83 | 0.90 | 0.727 |
| 0.122 | 0.71 | 0.95 | 0.659 |
| 0.189 | 0.18 | 0.99 | 0.164 |

**Figure 4.**
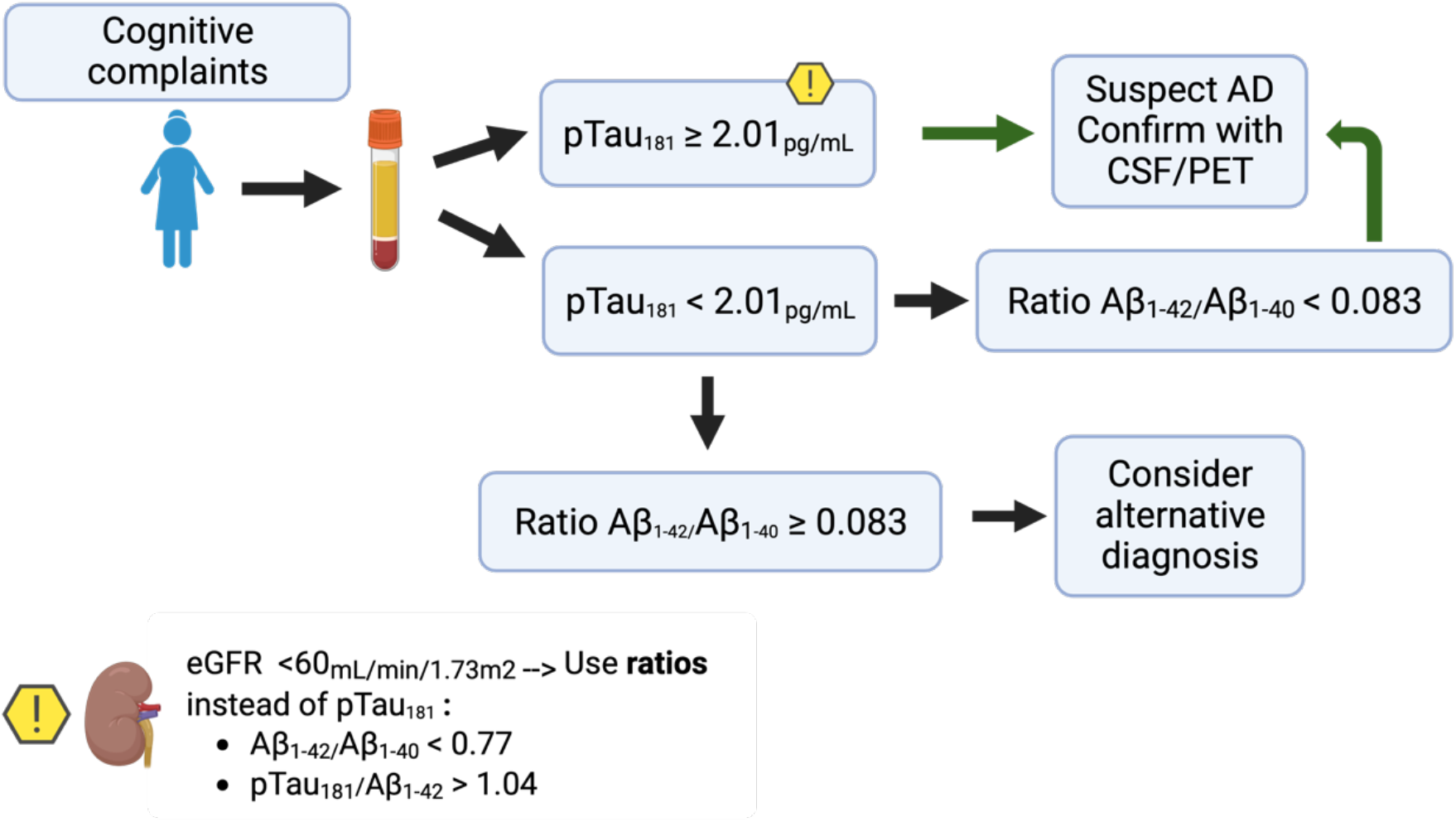
Algorithm of possible implementation of plasma markers in the evaluation of patients with cognitive complaints. pTau_181_: phosphorylated tau 181. Aβ_1–42_: Amyloid β_1–42_. Aβ_1–40_: Amyloid β_1–40_. eGFR: estimated glomerular filtration rate.

### Change in diagnostic accuracy of plasma biomarkers in specific clinical situations

We also tested the diagnostic performance of a cutoff value of 2.01pg/mL for plasma pTau_181_ discriminating A+T+ from A-T- in different subsets of our cohort considering the variables that were associated with plasma biomarkers. We tested the accuracy of this cutoff separately in Down syndrome and without Down syndrome, in the participants with dementia and without dementia, and in different stages of chronic kidney disease. We found that the accuracy was similar in all subgroups except for participants with chronic kidney disease (CKD) (**Supplementary Table**).

In the group with CKD 3a (eGFR <60mL/min/1.72m^2^), this cutoff had the lowest sensitivity (90%). While the false negative rate was still low (10%), a high false positive rate was observed (91%), suggesting that this cutoff would not be useful in this specific population. As we found that ratios did not differ significantly between CKD stages 1 to 3a, we tested whether the use of ratios could improve the accuracy in the group with CKD 3a. Using eGFR adapted cutoffs for Aβ_1–42_/Aβ_1–40_ or pTau_181_/Aβ_1–42_ in the subset of participants with CKD stage 3a reduced the false positive rate compared to pTau_181_ alone (27% to 9% of false positive, respectively) with the same 10% false negative rate (**Supplementary Material**).

## Discussion

In this study, we found that the concentration of plasma pTau_181_, and the ratios Aβ_1– 42_/Aβ_1–40_ and pTau_181_/Aβ_1–42_ measured in an automated platform, yielded good accuracy to detect the AD pathophysiology. These plasma biomarkers also showed moderate to high correlation with their CSF counterparts.

In our study the highest diagnostic yield was obtained by analyzing pTau_181_ first, followed by the Aβ_1–42_/Aβ_1–40_ ratio maximizing the number of A+T+ patients, while maintaining an acceptable false positive rate. Participants with positive plasma biomarkers would require confirmation with the current diagnostic gold standard (CSF or amyloid PET). In patients with advanced chronic kidney disease, the use of ratios could reduce the impact of having higher plasma concentrations associated to low renal function, thus minimizing the false positive rates in this population.

The performance of plasma markers to discriminate patients with AD from cognitively unimpaired participants, patients with other dementias and with not degenerative dementias has been assessed in previous studies using other analytical platforms, with AUCs ranging from 0.70 to 0.96 for pTau_181_[7,17,23–27], and from 0.64 to 0.86 for Aβ_1– 42_/Aβ_1–40_ [5,8,10]. Most of the studies reported better accuracies with the use of composite measures that combined two or more markers and/or clinical or genetic information. In our study, plasma pTau_181_ and the Aβ_1–42_/Aβ_1–40_ ratio measured with a fully automated platform showed high diagnostic performance to detect patients with AD CSF pathophysiology, and composite measures did not perform significantly better than pTau_181_ alone.

The accuracy of pTau_181_ in plasma has been previously studied in the Lumipulse and different accuracies have been reported to differentiate CU from AD. While Janelidze et al. reported an AUC of 0.7 [17] Wilson et al. found an accuracy of 0.96[24]. In our study, we found a global accuracy of 0.91 for pTau_181_. Different reasons could explain the differences between/across studies, including preanalytical conditions[28], characteristics of the sample and cohort, and the design of the study.

The effect of comorbidities as CKD on plasma biomarker concentrations points in the same direction as recently published studies[29] in which the use of ratios, in this case different isoforms of pTau with their corresponding unphosphorylated peptides, even analyzed on different platforms, could attenuate the effect of CKD.

The implications of implementing plasma biomarkers in primary care centers remain to be defined. While it has the potential to enhance the identification of patients at risk of neurodegenerative diseases, the possible consequences of positive results in asymptomatic individuals, as well as the risk of false positives, must be considered[4,18].

The strengths of our study are that we included all consecutive participants assessed throughout a year in our memory clinic with a variety of clinical diagnoses. This approach reduces the possibility of biases and ensures a reliable representation of the population assessed in the setting of a specialized memory clinic. In addition, the fact that all participants had CSF biomarkers, allowed us to compare plasma measures with their counterparts in CSF measured in the same analytical platform. Other strengths in our study are the fact that all markers were measured using the same batch of reagents and that the clinical information available allowed us to analyze the potential impact of comorbidities. Our study also has some limitations. Although we analyzed samples from non-selected consecutive participants, the criteria required that all participants had CSF. Therefore, the extrapolation to other contexts of use different than specialized memory units, such as primary care or population screening programs, should be made cautiously. Another limitation is the lack of Amyloid/Tau PET or neuropathological confirmation in our participants.

In summary, our study provides evidence that plasma markers can reliably be measured in an automated platform, showing great potential for the detection of AD pathophysiology in the context of a memory clinic. With the upcoming arrival of disease-modifying treatments into clinical practice, it is urgent to have screening methods to efficiently identify patients candidates to treatment. Although the results of the study need to be validated in multicenter studies, our findings suggest that plasma biomarkers measured with a fully automated platform could be integrated in specialized centers as a tool to screen patients with cognitive complaints at risk of having AD, and the diagnosis then confirmed using CSF. This could help to accelerate diagnosis and access to disease modifying therapies.

## Data Availability

Raw anonymized data and code for statistical analysis are available upon reasonable request. All requests should be sent to the corresponding author detailing the study hypothesis and statistical analysis plan. The steering committee of this study will decide whether data/code sharing is appropriate based on the novelty and scientific rigor of the proposal. All applicants will be asked to sign a data access agreement.

