## Supplementary material for "Diagnostic performance of plasma Aβ_1-42_, Aβ_1-40_ and pTau_181_ in the LUMIPULSE automated platform for the detection of Alzheimer disease"

### Supplementary Table 1: Demographics according to AT status

Unless otherwise specified, quantitative measures are presented as mean (SD).

$pTau_{181}$, phosphorylated tau 181. $A\beta_{1-42}$, Amyloid $\beta_{1-42}$. $A\beta_{1-40}$, Amyloid $\beta_{1-40}$VRF, vascular risk factors. CKD, Chronic kidney disease, DLP, dyslipidemia, HBP, high blood pressure. DM, diabetes mellitus. CU, cognitively unimpaired. OtherDem, other dementias. OtherNotDeg, not degenerative dementias. Down syndrome.

| | A-T- | A-T+ | A+T- | A+T+ |
|---|---|---|---|---|
| | 173 | 13 | 39 | 142 |
| Age | 60.2 (15.4) | 70.7 (10.5) | 68.1 (13) | 69.2 (11.4) |
| MMSE score | 26.6 (4.05) | 24.3 (3.74) | 23.7 (4.68) | 23 (4.21) |
| CSF $A\beta_{1-42}$ (pg/mL) | 978 (391) | 1216 (524) | 457 (151) | 544 (187) |
| CSF AB40 (pg/mL) | 9981 (3479) | 15213 (5414) | 8760 (2867) | 12488 (4528) |
| CSF $A\beta_{1-42}$/ $A\beta_{1-40}$ | 0.097 (0.0134) | 0.0789 (0.0145) | 0.0528 (0.0073) | 0.0446 (0.0085) |
| CSF tTau (pg/mL) | 278 (139) | 601 (161) | 312 (70.1) | 833 (402) |
| CSF $pTau_{181}$ (pg/mL) | 33.5 (12.2) | 81.8 (8.6) | 46.1 (10.2) | 139 (86.5) |
| Plasma $pTau_{181}$ (pg/mL) | 1.98 (0.844) | 2.72 (1.31) | 3 (0.99) | 3.9 (1.65) |
| plasma $A\beta_{1-42}$ (pg/mL) | 27.3 (6.99) | 29.2 (11.9) | 26.6 (6.78) | 26.3 (7.28) |
| plasma $A\beta_{1-40}$ (pg/mL) | 325 (80.1) | 373 (137) | 364 (74.9) | 362 (94.4) |
| plasma $A\beta_{1-42}$/ $A\beta_{1-40}$ | 0.0842 (0.0087) | 0.078 (0.006) | 0.0725 (0.0066) | 0.0728 (0.0065) |
| plasma $pTau_{181}$/$A\beta_{1-42}$ | 0.0765 (0.0464) | 0.0932 (0.0283) | 0.115 (0.0329) | 0.151 (0.0642) |
| Female % | 56.6 | 61.5 | 56.4 | 58.5 |
| *APOE4+* % | 14 | 30.8 | 41 | 38.7 |
| A+T+ % | 0 | 0 | 0 | 100 |
| VRF % | 63.1 | 90.9 | 69.4 | 69.8 |
| HBP % | 54 | 90.9 | 63.3 | 56 |
| DLP % | 47.1 | 80 | 46.7 | 59.8 |
| DM % | 18 | 60 | 10 | 28.2 |
| CKD ≥3a % | 6.88 | 15.4 | 7.69 | 7.09 |
| Stroke % | 14 | 0 | 18.5 | 5.26 |
| CPAP % | 9.43 | 16.7 | 3.45 | 9.52 |
| CU/AD/OtherNotDeg/ OtherDem/Down/Uncertain | 55/0/49/15/23 /31 | 1/0/1/3/1/7 | 0/2/7/8/9/13 | 1/95/4/6/32/4 |
| A-T-/A-T+/A+T-/A+T+ | 173/0/0/0 | 0/13/0/0 | 0/0/39/0 | 0/0/0/142 |

**Supplementary Table 2: Accuracy of Aβ$_{1-42}$/ Aβ$_{1-40}$ to differentiate A+T+ de A-T- in those patients with pTaup$_{181}$ <2.01 pg/mL (pTau$_{181}$ negative)**

| Threshold Aβ$_{1-42}$/ Aβ$_{1-40}$ in pTau$_{181}$ negative | Sensitivity | Specificity | Youden |
|---|---|---|---|
| 0.083 | 1.00 | 0.65 | 0.652 |
| 0.078 | 0.88 | 0.83 | 0.701 |
| 0.077 | 0.75 | 0.84 | 0.593 |
| 0.076 | 0.63 | 0.85 | 0.477 |
| 0.075 | 0.63 | 0.90 | 0.529 |
| 0.071 | 0.63 | 0.95 | 0.573 |
| 0.066 | 0.38 | 0.99 | 0.366 |

### Supplementary Table 3: Diagnostic performance of pTau$_{181}$ in plasma in different subsets with a cutoff value of 2.01 pg/mL (Sensitivity 95% in total sample)

pTau$_{181}$, phosphorylated tau 181. eGFR, estimated glomerular filtration rate. PPV, positive predictive value. NPV, negative predictive value.

| pTau$_{181}$ >2,01 pg/mL | Sensitivity | Specificity | PPV | NPV |
|---|---|---|---|---|
| Total | 95% | 65% | 69% | 93% |
| Euploid | 94% | 65% | 66% | 93% |
| Down syndrome | 100% | 65% | 80% | 100% |
| Dementia | 96% | 65% | 66% | 93% |
| Not Dementia | 94% | 68% | 58% | 96% |
| eGFR>90 | 92% | 76% | 52% | 97% |
| eGFR 60-90 | 96% | 58% | 79% | 90% |
| eGFR<60* | 90% | 9% | 47% | 50% |

Supplementary material. *Diagnostic performance of plasma $A\beta_{1-42}$, $A\beta_{1-40}$ and $pTau_{181}$ in the LUMIPULSE automated platform for the detection of Alzheimer disease*. J. Arranz et al. 2023.

**Supplementary Table 4 : Performance of plasma $pTau_{181}$ cutoff selected for total sample in different CKD subsets (stage 1 to 3a), compared to plasma $pTau_{181}$ cutoff obtained from different CKD subsets**

CKD, Chronic kidney disease. Obtained from whole sample: accuracy with our selected cutoff values for total sample applied to the specified range of eGFR. Obtained from population with CKD: accuracy of biomarkers adjusted by renal function with a pre-established sensitivity of 95%.

| | | $pTau_{181}$ | | | | $A\beta_{1-42}/A\beta_{1-40}$ | | | | $pTau_{181}/A\beta_{1-42}$ | | | |
|---|---|---|---|---|---|---|---|---|---|---|---|---|---|
| | | Cutoff | Sens | Spec | Youden | Cutoff | Sens | Spec | Youden | Cutoff | Sens | Spec | Youden |
| CKD stage 1 | Obtained from whole sample | 2.01 | 0.91 | 0.77 | 0.681 | 0.083 | 0.92 | 0.66 | 0.575 | 0.075 | 0.83 | 0.68 | 0.516 |
| | Obtained from population with CKD stage 1 | 1.57 | 0.96 | 0.41 | 0.370 | 0.083 | 0.96 | 0.62 | 0.582 | 0.068 | 0.96 | 0.58 | 0.535 |
| CKD stage 2 | Obtained from whole sample | 2.01 | 0.96 | 0.58 | 0.541 | 0.083 | 0.94 | 0.55 | 0.481 | 0.075 | 0.97 | 0.59 | 0.566 |
| | Obtained from population with CKD stage 2 | 2.1 | 0.95 | 0.66 | 0.610 | 0.083 | 0.95 | 0.53 | 0.485 | 0.088 | 0.95 | 0.86 | 0.812 |
| CKD stage 3 | Obtained from whole sample | 2.01 | 0.9 | 0.09 | 0 | 0.083 | 1 | 0.36 | 0.264 | 0.075 | 0.9 | 0.36 | 0.263 |
| | Obtained from population with CKD stage 3 | 2.78 | 0.9 | 0.45 | 0.355 | 0.077 | 0.9 | 0.73 | 0.627 | 0.104 | 0.9 | 0.91 | 0.809 |

### Supplementary Table 5: Diagnostic accuracy of plasma biomarkers for the discrimination of A-T- from A+T+ in different subsets.

We only included biomarkers and models that yielded AUC >0.80. Shaded cells indicate AUC significantly different from model 0 (DeLong test, $p<0.05$, multiple comparisons). Asterisks indicate those AUC significantly higher than $pTau_{181}$.

CU, cognitively unimpaired; AD, Alzheimer Disease; OtherNotDeg, Not-neurodegenerative dementias. OtherDem, Other dementias. Model 0, Age+Sex+*APOE*4. Model 1, $pTau_{181}$ + $A\beta_{1-42}$/ $A\beta_{1-40}$. Model 2, $pTau_{181}$ + $A\beta_{1-42}$, Model 3, Age+Sex+*APOE*4+$pTau_{181}$+ $A\beta_{1-42}$+ $A\beta_{1-40}$.

| A-T- vs A+T+ (AUC 95% CI) | Total sample | Euploid | Triploid | Without dementia | Dementia | CU and AD | OtherDem and AD | OtherNotDeg and AD |
|---|---|---|---|---|---|---|---|---|
| $pTau_{181}$ | 0.91 (0.87-0.94) | 0.89 (0.85-0.93) | 0.95 (0.89-1) | 0.90 (0.85-0.94) | 0.87 (0.78-0.96) | 0.95 (0.92-0.98) | 0.85 (0.76-0.95) | 0.89 (0.83-0.95) |
| $A\beta_{1-42}$/ $A\beta_{1-40}$ | 0.86 (0.82-0.9) | 0.88 (0.84-0.92) | 0.81 (0.68-0.94) | 0.87 (0.83-0.92) | 0.79 (0.68-0.91) | 0.94 (0.91-0.98) | 0.81 (0.64-0.97) | 0.88 (0.82-0.95) |
| $pTau_{181}/A\beta_{1-42}$ | 0.92 (0.89-0.95) | 0.93* (0.9-0.96) | 0.94 (0.86-1) | 0.91 (0.87-0.95) | 0.88 (0.79-0.97) | 0.97 (0.95-1) | 0.91 (0.84-0.99) | 0.94 (0.9-0.99) |
| Model 0 | 0.73 (0.67-0.79) | 0.85 (0.8-0.89) | 0.93 (0.86-1) | 0.75 (0.67-0.82) | 0.68 0.55-0.8) | 0.94 (0.9-0.98) | 0.79 (0.68-9) | 0.83 (0.77-0.9) |
| Model 1 | 0.94 (0.92-0.97) | 0.95* (0.92-0.97) | 0.96 (0.9-1) | 0.94 (0.91-0.97) | 0.92 (0.85-1) | 0.99 (0.97-1) | 0.92 (0.85-0.99) | 0.94 (0.89-0.99) |
| Model 2 | 0.93 (0.9-0.96) | 0.93* (0.9-0.96) | 0.95 (0.89-1) | 0.92 (0.88-0.96) | 0.90 (0.81-0.98) | 0.97 (0.95-1) | 0.91 (0.83-1) | 0.95 (0.9-0.99) |
| Model 3 | 0.95* (0.93-0.98) | 0.96* (0.94-0.99) | 0.98 (0.94-1) | 0.95 (0.92-0.98) | 0.95 (0.91-0.99) | 1* (0.99-1) | 0.98 (0.95-1) | 0.98* (0.95-1) |

### Supplementary Table 6: Diagnostic accuracy of plasma biomarkers for the discrimination of A-T- and A+T+ from intermediate states (A-T+ and A+T-)

Shaded cells indicate AUC significantly different from model 0 (DeLong test, $p<0.05$, multiple comparisons). Asterisks indicate those AUC significantly higher than $pTau_{181}$.

Model 0, Age+Sex+*APOE*4. Model 1, $pTau_{181}$ + $A\beta_{1–42}$/ $A\beta_{1-40}$. Model 2, $pTau_{181}$ + $A\beta_{1–42}$, Model 3, Age+Sex+*APOE*4+$pTau_{181}$+ $A\beta_{1–42}$+ $A\beta_{1–40}$.

| Intermediate states (AUC 95% CI) | A-T- and A-T+ | A-T- and A+T- | A+T+ and A-T+ | A+T+ and A+T- | A-T+ and A+T- |
|---|---|---|---|---|---|
| $pTau_{181}$ | 0.75 (0.59-0.92) | 0.84 (0.78-0.9) | 0.75 (0.59-92) | 0.69 (0.6-0.78) | 0.63 (0.43-0.84) |
| $A\beta_{1–42}$/ $A\beta_{1-40}$ | 0.73 (0.58-0.87) | 0.86 (0.8-0.92) | 0.73 (0.58-0.87) | 0.53 (0.43-0.64) | 0.74 (0.59-0.89) |
| $pTau_{181}$/$A\beta_{1–42}$ | 0.85 (0.75-0.96) | 0.86 (0.8-0.92) | 0.85 (0.75-0.96) | 0.72 (0.64-0.81) | 0.73 (0.57-0.89) |
| Model 0 | 0.57 (0.42-0.73) | 0.77 (0.68-0.86) | 0.57 (0.42-73) | 0.49 (0.38-0.61) | 0.55 (0.39-0.72) |
| Model 1 | 0.85 (0.73-0.97) | 0.92 (0.88-0.96) | 0.85 (0.73-0.97) | 0.69 (0.6-0.78) | 0.79 (0.63-0.94) |
| Model 2 | 0.86 (0.75-0.96) | 0.87 (0.81-0.92) | 0.86 (0.75-0.96) | 0.73 (0.64-0.81) | 0.73 (0.57-0.89) |
| Model 3 | 0.92 (0.86-0.98) | 0.93* (0.93-0.97) | 0.92 (0.86-98) | 0.72 (0.64-0.81) | 0.85 (0.75-0.96) |

### Supplementary Table 7: Comparison of plasma biomarkers' accuracy using DeLong test adjusted by multiple comparisons

AD plasma biomarkers and their combinations that better discriminate A+T+ from A-T-. We included those with AUC >0.80 and $p<0.05$.

Model 0: Age+Sex+*APOE4*. Model 1, $pTau_{181}$ + $A\beta_{1-42}/A\beta_{1-40}$. Model 2, $pTau_{181}$ + $A\beta_{1-42}$. Model 3, Age+Sex+*APOE*4+$pTau_{181}$+ $A\beta_{1-42}$+ $A\beta_{1-40}$.

| Contrast | BM1 | AUC1 | BM2 | AUC2 | pvalue | sig |
|---|---|---|---|---|---|---|
| A-T- vs. A+T+ | $pTau_{181}$ | 0.91 | $pTau_{181}/A\beta_{1-42}$ | 0.92 | 0.18226206 | ns |
| A-T- vs. A+T+ | $pTau_{181}$ | 0.91 | $A\beta_{1-42}/A\beta_{1-40}$ | 0.86 | 0.07643099 | ns |
| A-T- vs. A+T+ | $pTau_{181}$ | 0.91 | Model 0 | 0.73 | 6.2397E-07 | <0.05 (comp.mult) |
| A-T- vs. A+T+ | $pTau_{181}$ | 0.91 | Model 1 | 0.94 | 0.00245538 | ns |
| A-T- vs. A+T+ | $pTau_{181}$ | 0.91 | Model 2 | 0.93 | 0.00673938 | ns |
| A-T- vs. A+T+ | $pTau_{181}$ | 0.91 | Model 3 | 0.95 | 0.0006526 | <0.05 (comp.mult) |
| A-T- vs. A+T+ | $pTau_{181}/A\beta_{1-42}$ | 0.92 | $A\beta_{1-42}/A\beta_{1-40}$ | 0.86 | 0.00758168 | ns |
| A-T- vs. A+T+ | $pTau_{181}/A\beta_{1-42}$ | 0.92 | Model 0 | 0.73 | 6.8132E-10 | <0.05 (comp.mult) |
| A-T- vs. A+T+ | $pTau_{181}/A\beta_{1-42}$ | 0.92 | Model 1 | 0.94 | 0.04754348 | ns |
| A-T- vs. A+T+ | $pTau_{181}/A\beta_{1-42}$ | 0.92 | Model 2 | 0.93 | 0.15863615 | ns |
| A-T- vs. A+T+ | $pTau_{181}/A\beta_{1-42}$ | 0.92 | Model 3 | 0.95 | 0.00315677 | ns |
| A-T- vs. A+T+ | $A\beta_{1-42}/A\beta_{1-40}$ | 0.86 | Model 0 | 0.73 | 4.7903E-05 | <0.05 (comp.mult) |
| A-T- vs. A+T+ | $A\beta_{1-42}/A\beta_{1-40}$ | 0.86 | Model 1 | 0.94 | 4.8444E-06 | <0.05 (comp.mult) |
| A-T- vs. A+T+ | $A\beta_{1-42}/A\beta_{1-40}$ | 0.86 | Model 2 | 0.93 | 0.00401329 | ns |
| A-T- vs. A+T+ | $A\beta_{1-42}/A\beta_{1-40}$ | 0.86 | Model 3 | 0.95 | 7.111E-07 | <0.05 (comp.mult) |
| A-T- vs. A+T+ | Model 0 | 0.73 | Model 1 | 0.94 | 5.0997E-11 | <0.05 (comp.mult) |
| A-T- vs. A+T+ | Model 0 | 0.73 | Model 2 | 0.93 | 9.6752E-10 | <0.05 (comp.mult) |
| A-T- vs. A+T+ | Model 0 | 0.73 | Model 3 | 0.95 | 1.0208E-13 | <0.05 (comp.mult) |
| A-T- vs. A+T+ | Model 1 | 0.94 | Model 2 | 0.93 | 0.10695235 | ns |
| A-T- vs. A+T+ | Model 1 | 0.94 | Model 3 | 0.95 | 0.068173 | ns |
| A-T- vs. A+T+ | Model 2 | 0.93 | Model 3 | 0.95 | 0.0134703 | ns |

Supplementary material. *Diagnostic performance of plasma $A\beta_{1-42}$, $A\beta_{1-40}$ and $pTau_{181}$ in the LUMIPULSE automated platform for the detection of Alzheimer disease*. J. Arranz et al. 2023.

### Supplementary Figure 1: Correlation maps by cognitive status.

$pTau_{181}$, phosphorylated tau 181. $A\beta_{1-42}$, Amyloid $\beta_{1-42}$. $A\beta_{1-40}$, Amyloid $\beta_{1-40}$

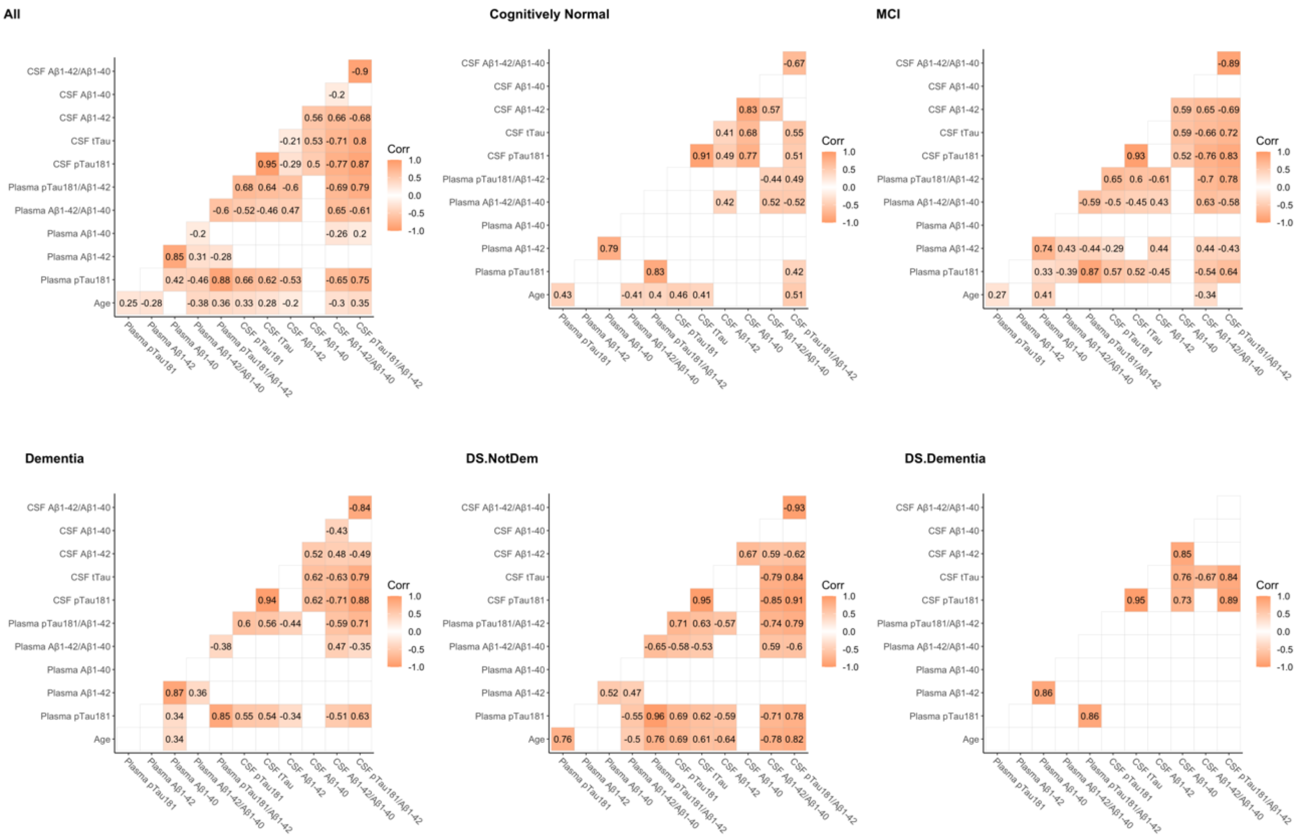

**Supplementary Figure 2: Diagnostic accuracy of plasma markers to detect the Alzheimer's disease pathophysiological profile.**

pTau$_{181}$, phosphorylated tau 181. Aβ$_{1–42}$, Amyloid β$_{1–42}$. Aβ$_{1–40}$, Amyloid β$_{1–40}$.

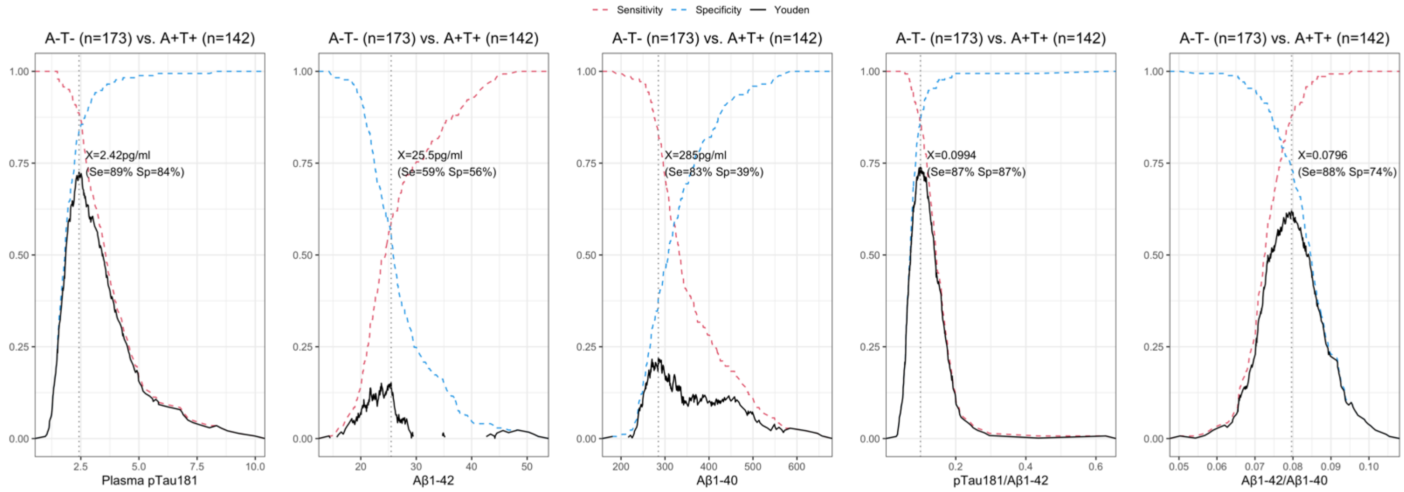

Supplementary material. *Diagnostic performance of plasma Aβ$_{1-42}$, Aβ$_{1-40}$ and pTau$_{181}$ in the LUMIPULSE automated platform for the detection of Alzheimer disease*. J. Arranz et al. 2023.

### Supplementary Figure 3: Areas Under the ROC Curve of plasma markers to detect Alzheimer's disease

CU, cognitively unimpaired; AD, Alzheimer Disease; OtherDem, Other dementias; MCI, mild cognitive impairment. pTau$_{181}$, phosphorylated tau 181. Aβ$_{1–42}$, Amyloid β$_{1–42}$. Aβ$_{1–40}$, Amyloid β$_{1–40}$. Model 0, Age+Sex+*APOE*4. Model 1, pTau$_{181}$ + Aβ$_{1–42/}$ Aβ $_{1-40}$. Model 2, pTau$_{181}$ + Aβ$_{1–42}$, Model 3, Age+Sex+*APOE*+pTau$_{181}$+ Aβ$_{1–42}$+ Aβ$_{1–40}$.

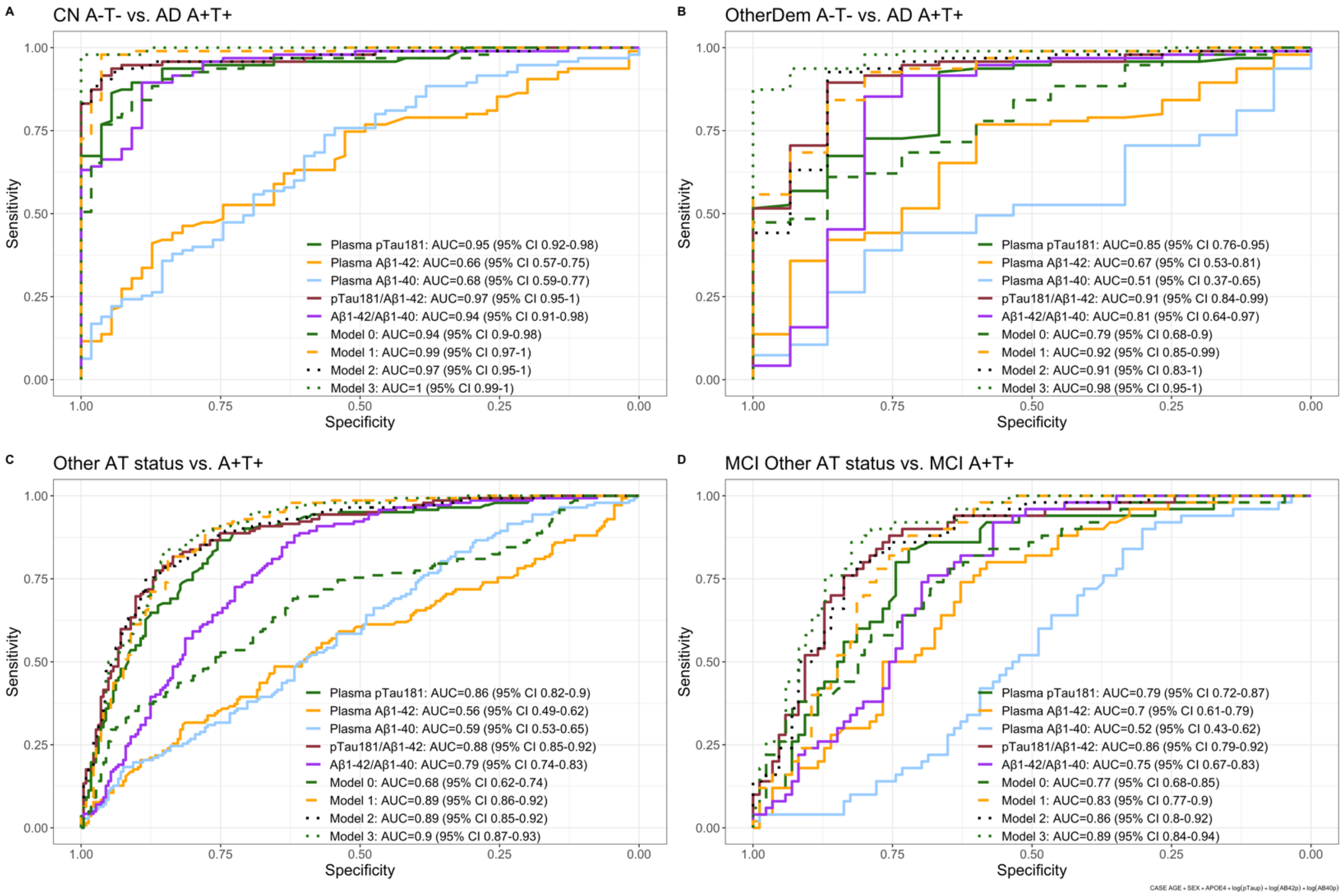
